# Nirsevimab effectiveness, number needed to immunize and impact on severe RSV outcomes in preterm, high-risk and healthy-term infants, Quebec, Canada

**DOI:** 10.1101/2025.07.27.25332262

**Authors:** Sara Carazo, Manale Ouakki, Danuta M Skowronski, Maude Paquette, Nicholas Brousseau, Denis Talbot, Charles-Antoine Guay, Caroline Quach, Rodica Gilca, Jesse Papenburg

## Abstract

**Background:** During the 2024-25 season, a universal infant nirsevimab program was publicly funded in Quebec, Canada. We estimated effectiveness, number-needed-to-immunize (NNI) and impact against severe respiratory syncytial virus (RSV) outcomes.

**Methods:** We conducted a test-negative study among nirsevimab-eligible children RSV-tested during ER consultation or hospitalization between October 1^st^, 2024 and March 3^st^, 2025. Eligible children were healthy-term and born during the RSV season (at-birth group) or <6 months old on October 1^st^ (catch-up group), preterm, with high-risk conditions or living in remote regions. We estimated adjusted effectiveness by eligible group and further used 2023-24 respiratory hospitalization rates, RSV-positivity from a systematic hospital-based surveillance network, and coverage estimates to derive the NNI and tally of averted hospitalizations and ICU admissions.

**Results:** Effectiveness analyses included 3,172 ER consultations (668 RSV-positive) and 1,758 hospitalizations (549 RSV-positive). Nirsevimab effectiveness against ER consultation, hospitalization and ICU admission was 86% (95%CI:82-90), 89% (95%CI:84-92) and 88% (95%CI:58-97), respectively. Effectiveness exceeded 80% for all eligible groups. We estimate 41 at-birth and 58 catch-up immunizations needed to avert one RSV-associated hospitalization between October and March. Applying Quebec catch-up coverage (64%) and timing (November launch), we estimate more than half of RSV-associated hospitalizations and ICU admissions were prevented, potentially increasing to more than three-quarters if catch-up coverage reached 90% by October 1^st^.

**Conclusions:** Nirsevimab is highly effective and could more substantially impact the overall burden of RSV hospitalization and ICU admission through broad and timely administration to healthy-term and high-risk infants. To inform optimal nirsevimab timing, the durability of late-season protection warrants further investigation.

## INTRODUCTION

Respiratory syncytial virus (RSV) is a leading cause of emergency room (ER) visits and hospitalizations among young children in North America, with the highest hospitalization risk during the first three months of life [1]. While underlying health conditions predispose children to severe RSV illness, most pediatric RSV hospitalizations occur in healthy full-term infants [2].

Nirsevimab is a long-acting monoclonal antibody, which, in clinical trials, reduced the risk of medically attended lower respiratory tract infection and hospitalization due to RSV by approximately 80% for 150 days [3,4]. Similarly substantial effectiveness (75%-83%) was reported in a 2025 systematic review of studies of nirsevimab programs implemented during the 2023-24 RSV season in Europe and the USA [5]. However, there are no data on effectiveness among preterm infants and high-risk populations, and evidence on health systems impact (e.g., hospitalizations averted) and number needed to immunize (NNI) remain sparse [6,7].

Nirsevimab was first authorized in Canada on April 19, 2023 [8]. In May 2024, Canada’s National Advisory Committee on Immunization recommended building towards a universal RSV immunization program for all infants during their first RSV season [9]. The province of Quebec, with a population of 9 million and about 86,000 births annually, became the first in Canada to announce a publicly-funded universal infant nirsevimab program with all other provinces and territories remaining undecided [10–12]. By the fall of 2024, Quebec and Ontario were the only two Canadian provinces to implement such a program for the 2024-25 season [10,13].

This study among Quebec infants eligible for nirsevimab immunization during the 2024-25 season aimed to estimate: effectiveness of nirsevimab against RSV-associated ER consultations, hospitalizations and intensive care unit (ICU) admissions, overall, by eligible group and by time since immunization; and NNI and tally of hospitalizations and ICU admissions averted.

## METHODS

### Study design and population

A test-negative case-control study evaluated nirsevimab effectiveness against: RSV-associated ER consultations, hospitalizations and ICU admissions. The study population included all Quebec children eligible for nirsevimab immunization who had a unique identifier number (allowing dataset merging), who were assessed for RSV by nucleic acid amplification test (NAAT) from October 1, 2024, to March 31, 2025, and who had an ER consultation or a hospitalization between 7 days before and 2 days after testing. All children born during the 2024-25 RSV season were eligible as were the following groups in relation to age at the start of the RSV season: (1) healthy full-term children <6 months old; (2) children born prematurely <8 months old; (3) children living in remote regions, such as *Nunavik* and *Terres-Cries-de-la-Baie-James* <8 months old; and (4) children with one of the following chronic conditions increasing the risk of severe RSV disease <19 months old: bronchopulmonary dysplasia, chronic lung disease, congenital heart disease or cardiomyopathy, pulmonary hypertension, Down syndrome, cystic fibrosis, neuromuscular disorder or congenital anomaly that impairs clearance of upper airway secretions, and bone marrow, stem cell, or solid organ transplantation [10]. Comorbidities were identified using ICD-10 codes specified in the main or secondary diagnosis of any hospitalization or ER consultation since birth (Supplementary_Table_1). Prematurity was identified by the encoded duration of pregnancy in the birth hospitalization and the ICD-10 code P07.3. RSVpreF vaccine was authorized for use in pregnancy in Canada in January 2024 but was not publicly funded in Quebec during the 2024-25 season [14]; regardless, we excluded any children born to mothers vaccinated with RSVpreF vaccine. Considering the RSV incubation period and delay between symptom onset and consultation, we also excluded children who received nirsevimab <7 days before consultation or admission. In NNI analyses, we included children born in Quebec during the 2023-24 RSV season (from October to March) or <6 months of age on October 1^st^, 2023.

### Data sources

For nirsevimab effectiveness analyses we used a unique personal identifier to combine four administrative databases: the Quebec provincial immunization registry, including all Quebec residents and all administered immunizations; the Quebec sentinel laboratory registry, with approximately 80 laboratories in the province of Quebec contributing NAAT results; the common ER database; and the administrative hospitalization database. The ER and hospitalization databases contain administrative and clinical information, including reason for ER consultation and ICD-10 codes for discharge diagnosis. By the May 30, 2025, data extraction date, information on discharge diagnosis was complete for 98% of ER consultations and 75% of hospitalizations.

For NNI analyses, we used: the administrative hospitalization database; the Quebec sentinel surveillance network of six hospitals that conduct systematic testing of hospital admissions for acute respiratory infection using a multiplex PCR panel for respiratory viruses [15]; and birth data from the *Institut de la statistique du Quebec* [16].

### Exposure and outcome definitions

Immunized children received nirsevimab ≥7 days before ER consultation or hospitalization.

ER consultations were NAAT-positive for RSV between 7 days before and 2 days after consultation not followed by hospital admission. Hospitalizations and ICU admissions were similarly NAAT-positive for RSV with hospital admission lasting >24 hours. We performed sensitivity analyses restricted to NAAT-positive RSV consultations or hospitalizations with any respiratory presentation, or for bronchiolitis or bronchitis, as reasons for ER consultation or as the main discharge diagnosis, as specified in Supplementary_Table_2.

### Effectiveness analyses

For each outcome, logistic regression models compared the odds of immunization between test-positive cases and test-negative controls. Models were adjusted for sex, age in months at testing, region of residence, social and material deprivation indices, presence of comorbidities (dichotomous variable), prematurity, and two-week periods of admission. Each child could contribute several ER consultations or hospitalizations during the study period if associated with a test for RSV but was censored after the first positive test. The same hospitalized test-negative controls were used to estimate effectiveness against hospitalization and ICU admission. Nirsevimab effectiveness (%) and 95% confidence intervals (95%CI) were calculated as (1 − *adjusted odds ratio*) × 100. Stratified analyses evaluated effectiveness separately for each eligible group, for infants born during the RSV season (at-birth group) or <6 months old in October 2024 (catch-up group), and by time (four-week intervals) since nirsevimab administration.

### Statistical analyses to estimate NNI and hospitalizations averted

We used administrative hospitalization data for the most recent 2023-24 RSV season, preceding the nirsevimab program in Quebec, to estimate the number of respiratory and bronchiolitis admissions, as previously defined, among all newborns and infants <6 months old in October 2023. Nirsevimab was not available in Québec before fall 2024. We inferred the number of RSV-attributable hospitalizations by applying the proportion of tests being RSV-positive (RSV positivity) among respiratory or bronchiolitis hospitalizations within the Quebec hospital-based surveillance network during the 2023-24 season by the specified age groups. The number of RSV-attributable ICU admissions were estimated using the proportion of ICU admissions among RSV-positive hospitalizations in the Quebec surveillance network. We calculated the RSV hospitalization rate (per 100,000 infants) for each age group and diagnosis as 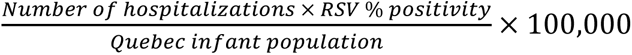 and NNI as 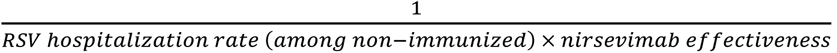, based on our nirsevimab effectiveness estimations and 95%CIs. We used the 2024-25 nirsevimab coverage data to estimate the number and proportion of averted hospitalizations among at-birth and catch-up groups considering catch-up immunization completed by October 1^st^, 2023, or, in sensitivity analyses, by November 1^st^, 2023. Similar calculations were used to estimate NNI to avert one ICU admission.

### Ethical considerations

The research protocol was examined by the Research Ethics Board of the Centre hospitalier universitaire de Québec-Université Laval and considered a program evaluation exempt from ethics review (request for advice number 2025-7907).

## RESULTS

### Population

During the study period, 13,306 specimens between October 1, 2023 and March 31, 2025, collected from all eligible age groups, were subjected to RSV NAAT. Of these, 3,172 RSV-tested specimens (from 2,966 children) were included in the estimation of nirsevimab effectiveness against ER consultations and 1,758 tests (from 1,583 children) against hospitalizations (Figure_1). The main reasons for exclusion were not being hospitalized or consulting at the ER (70.7% and 22.4%, respectively), age 6-18 months without eligibility risk factors (14.3% and 42.7%, respectively), and repeat tests performed after the initial testing for each hospitalization or ER consultation (1.9% and 1.9%, respectively).

**Figure 1:**
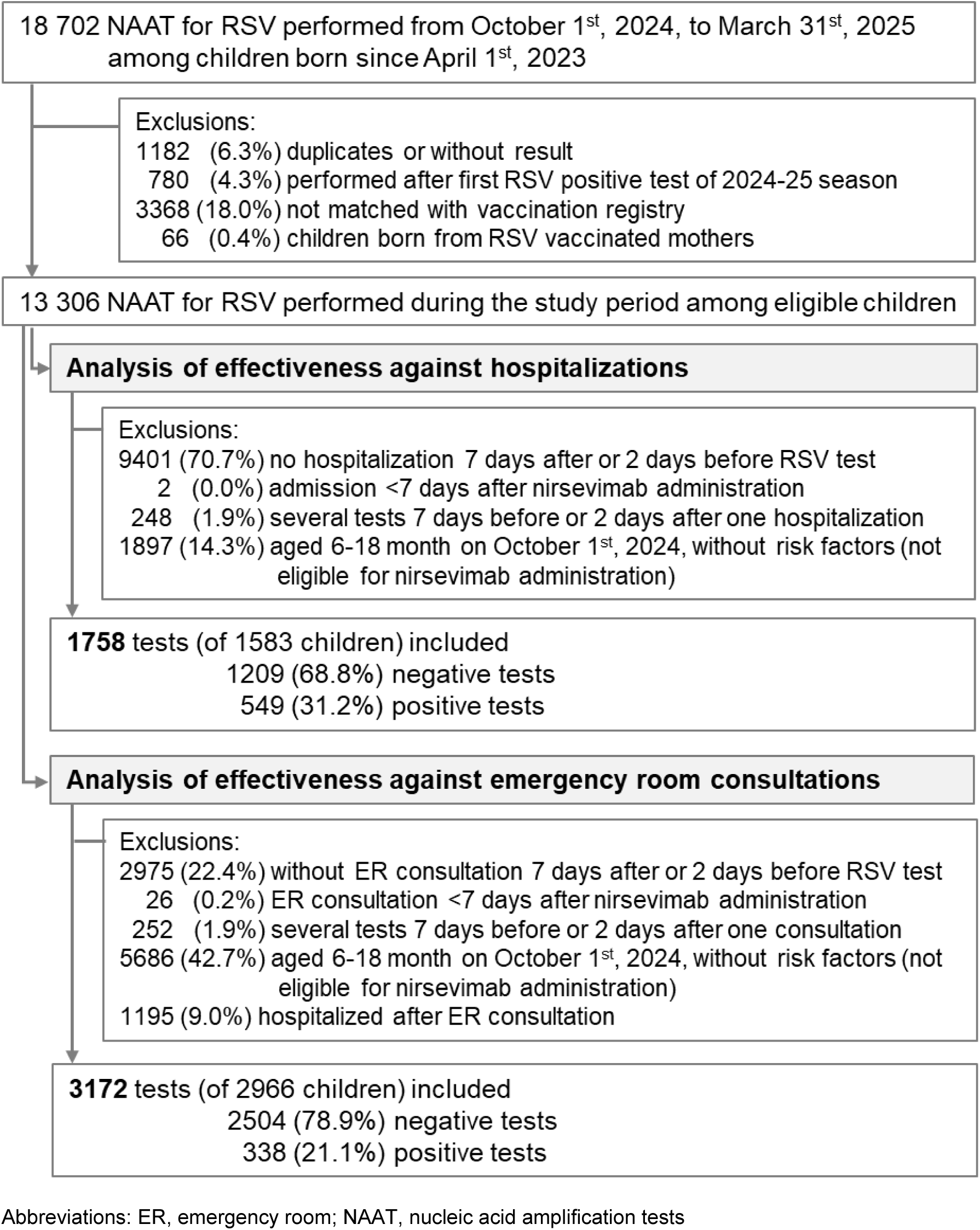
Population flowchart.

Among nirsevimab-eligible children during the 2024-25 season, RSV-positive NAAT included 668 ER consultations without hospitalization, 549 hospitalizations and 34 ICU admissions (Table_1). Most had a respiratory diagnosis (98%, 82% and 94%, respectively, for each outcome), while bronchiolitis/bronchitis was the main diagnosis for 70% of hospitalizations and 77% of ICU admissions. RSV-associated ER consultations and hospitalizations increased steadily starting in week 2024-40 and peaked at weeks 2024-46 to 2025-01, in line with Quebec sentinel laboratory network RSV positivity and with a similar profile in 2023-24 (Figure_2). The number of ER admissions and hospitalizations among infants from remote regions was insufficient to estimate nirsevimab effectiveness.

**Figure 2:**
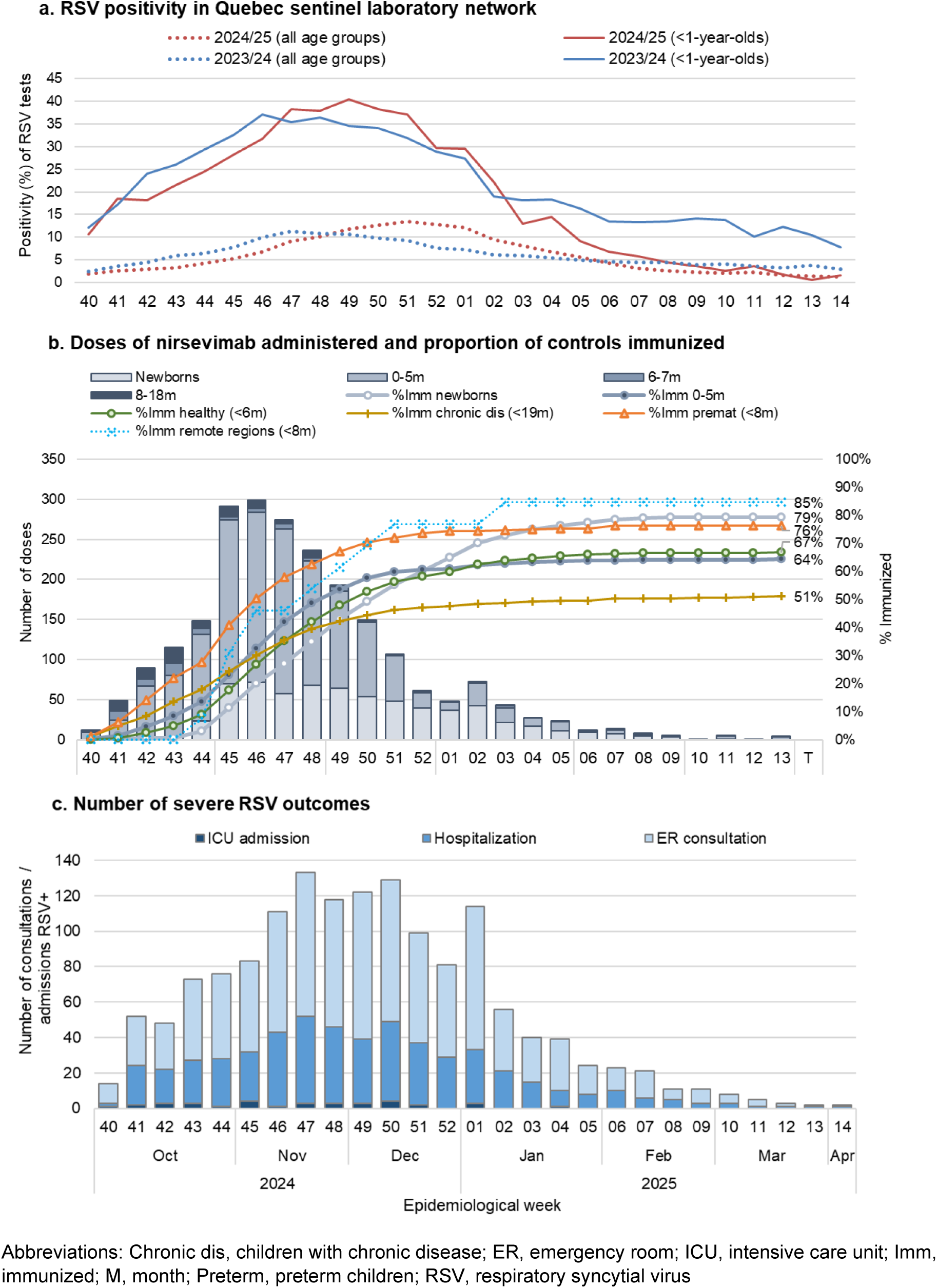
RSV circulation in Quebec, nirsevimab vaccination and RSV cases during the 2024-25 season.

Over half of participants were male and aged <6 months at RSV testing. Most ER consultations (71.3%, n=2339) and hospitalizations (59.8%, n=1052) occurred in healthy-term children (Table_1). While most participants belonged to the catch-up group, the at-birth group was more represented among controls (24.9% of ER consultations and 20.7% of hospitalizations) than RSV cases (7.6% and 8.7%, respectively).

**Table 1:** Characteristics of children included in the effectiveness analyses.

|  | Test-positive cases |  |  | Test-negative controls <sup>a</sup> |  |
| --- | --- | --- | --- | --- | --- |
|  | ER consultations | Hospitalizations | ICU admissions | ER consultations | Hospitalizations |
|  | N (%) | N (%) | N (%) | N (%) | N (%) |
| **N** | **668** | **549** | **34** | **2504** | **1209** |
| **Nirsevimab status** |  |  |  |  |  |
| Immunized | 69 (10.3) | 55 (10.0) | 3 (8.8) | 1265 (50.5) | 627 (51.9) |
| Days since immunization, median (IQR) | 53 (34 – 77) | 49 (31 – 76) | 41 (40-49) | 71 (49 – 98) | 67 (40 – 97) |
| **Sex** |  |  |  |  |  |
| Female | 293 (43.9) | 208 (37.9) | 12 (35.3) | 1104 (44.1) | 562 (46.5) |
| Male | 375 (56.1) | 341 (62.1) | 22 (64.7) | 1400 (55.9) | 647 (53.5) |
| **Age at testing, months** |  |  |  |  |  |
| Median (IQR) | 5.7 (3.3 – 7.5) | 4.8 (2.8 – 7.6) | 3.6 (2.1 – 6.0) | 5.4 (2.8 – 8.5) | 5.7 (3.1 – 9.1) |
| <6 months | 355 (53.1) | 339 (61.7) | 25 (73.5) | 1383 (55.2) | 623 (51.5) |
| 6-11 months | 263 (39.4) | 160 (29.1) | 5 (14.7) | 949 (37.9) | 420 (34.7) |
| 12-17 months | 47 (7.0) | 45 (8.2) | 3 (8.8) | 139 (5.6) | 124 (10.3) |
| 18+ months | 3 (0.4) | 5 (0.9) | 1 (2.9) | 33 (1.3) | 42 (3.5) |
| **Age on October 1<sup>st</sup>, 2024, months** |  |  |  |  |  |
| 0 (born after) | 51 (7.6) | 48 (8.7) | 1 (2.9) | 623 (24.9) | 250 (20.7) |
| 0-<6 months | 514 (76.9) | 406 (74.0) | 27 (79.4) | 1577 (63.0) | 706 (58.4) |
| 6-18 months | 103 (15.4) | 95 (17.3) | 6 (17.6) | 304 (12.1) | 253 (20.9) |
| **Delay test – admission, days, median (IQR)** | 0 (0 – 0) | 0 (-1 – 0) | 0 (-1 – 0) | 0 (0 – 0) | 0 (-1 – 0) |
| **Eligible groups, days since immunization <sup>b</sup>** |  |  |  |  |  |
| Healthy-term children | 476 (71.3) | 351 (63.9) | 23 (67.6) | 1863 (74.4) | 701 (58.0) |
| Days, median (IQR) | 54 (34 – 77) | 38 (23 – 74) | 40 (40 – 55) | 69 (49 – 94) | 64 (42 – 91) |
| Children with comorbidities | 135 (20.2) | 125 (22.8) | 9 (26.5) | 450 (18.0) | 402 (33.3) |
| Days, median (IQR) | 45 (16 – 70) | 52 (34 – 82) | 42 (42 – 42) | 75.5 (46 – 108) | 68 (35 – 103) |
| Preterm children | 65 (9.7) | 90 (16.4) | 4 (11.8) | 248 (9.9) | 184 (15.2) |
| Days, median (IQR) | 45 (42 – 55) | 50 (40.5 – 65.5) | NE | 81 (51 – 115) | 74 (38 – 112) |
| Children of remote regions | 1 (0.1) | 3 (0.5) | 0 (0.0) | 4 (0.2) | 16 (1.3) |
| Days, median (IQR) | NE | 163 (163 – 163) | NE | 43 (7 – 51) | 57 (22 – 73) |
| **Main diagnosis** |  |  |  |  |  |
| Missing | 9 (1.3) | 75 (13.7) | 0 (0.0) | 44 (1.8) | 432 (35.7) |
| Respiratory diagnosis <sup>c</sup> | 655 (98.1) | 451 (82.1) | 32 (94.1) | 2336 (93.3) | 491 (40.6) |
| Bronchiolitis / bronchitis diagnosis | 196 (29.3) | 383 (69.8) | 26 (76.5) | 169 (6.7) | 166 (13.7) |

<sup>a</sup> Test-negative hospitalizations were controls for both hospitalization and ICU effectiveness analyses
<sup>b</sup> Groups of children with comorbidities and preterm children are not mutually exclusive (100 hospitalizations and 139 ER consultations belong to both eligible subpopulations)
<sup>c</sup> For ER consultations, respiratory diagnosis is based on reason for consultation and discharge diagnosis
Abbreviations: ER, emergency room; ICU, intensive care unit; IQR, interquartile range; NE, not estimable
Note: Numbers are n and column percentages except otherwise specified

Due to a supply shortage in October 2024, only high-risk infants were prioritized that month. Most doses of nirsevimab were administered in November and December 2024 (Figure_2). Among test-negative children, the proportion immunized with nirsevimab by the end of the season was 79.2% for the at-birth group, 64.4% for the catch-up group, 66.8% for healthy-term children, 51.1% for children with chronic diseases and 76.3% for preterm children. Among ER consultations, 50.5% (n=1265) of controls vs. 10.3% (n=69) of cases were immunized with nirsevimab at least 7 days before testing (Table_1). Among hospitalizations, 51.9% (n=627) of controls vs. 10.0% (n=55) of cases were so immunized.

Median time since immunization was longer for high-risk than for healthy-term children (e.g. 38 days for healthy-term cases vs 52 days for children with chronic diseases).

### Nirsevimab effectiveness

Nirsevimab effectiveness overall was 86% (95%CI: 82-90) against ER consultation without hospitalization, 89% (95%CI: 84-92) against hospitalization and 88% (95%CI: 58-97) against ICU admission (Figure_3). Protection against ER consultations ranged 86% to 91% regardless of outcome definition (any, respiratory or bronchiolitis admission), age group or eligible group, except for slightly lower estimate of 83% (95%CI: 65-92) for the at-birth group and higher at 97% (95%CI: 91-99) for preterm children, albeit with overlapping CIs (Figure_3, Supplementary_Table 3). Effectiveness against hospitalizations ranged 89% to 93% for all groups except the at-birth group (84%; 95%CI: 58-94) and children with chronic diseases (79%; 95%CI:60-89) (Figure_3, Supplementary_Table 4). Effectiveness against hospitalizations was similar when restricted to respiratory diagnosis, suggesting limited misclassification when defining RSV-hospitalization only by a positive NAAT. Few ICU admissions precluded meaningful stratification (Supplementary_Table 5). Point estimates of effectiveness against ER consultations and hospitalizations were lower for those immunized ≥12 weeks prior, but with wide and overlapping 95%CIs following the drop in RSV infections in February 2025 (Figure_2, Figure_3). Trend tests of estimates stratified by time since immunization were not significant for any of the outcomes (Figure_3).

**Figure 3:**
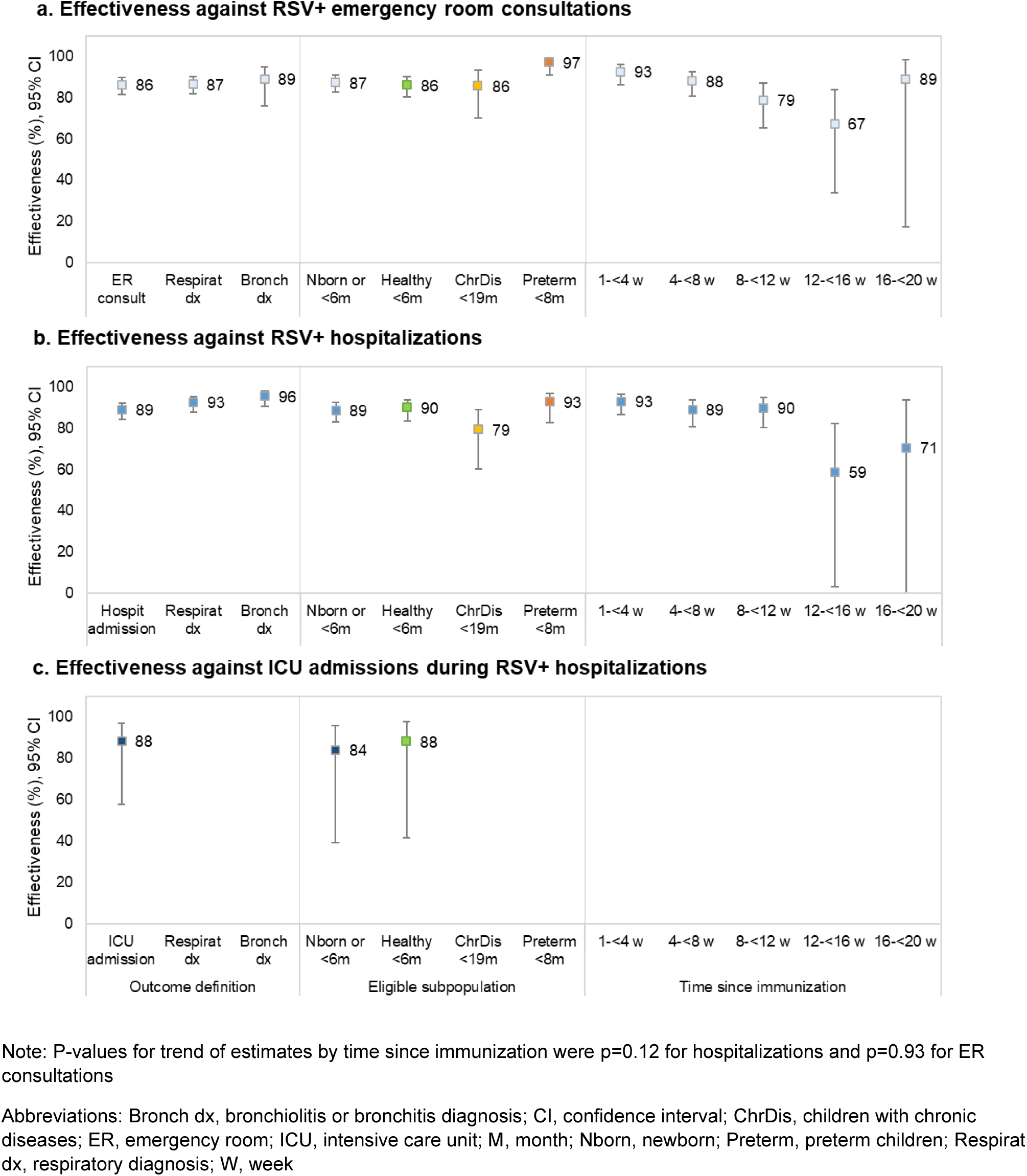
Nirsevimab effectiveness against severe outcomes by outcome definition, eligible subpopulation and time since immunization.

### NNI and averted hospitalizations/ICU admissions

Based on the 2023-24 RSV season, between 41 and 57 nirsevimab at-birth immunizations would be required to avert one RSV hospitalization or one RSV hospitalization with bronchiolitis/bronchitis diagnosis, respectively (Table_2). Among the catch-up group, corresponding NNIs were 58 and 63 if immunized by 1st October. Assuming nirsevimab coverage of 79% and 64% within the at-birth and catch-up groups, respectively (as per the 2024-25 season), we estimate 742 (73%) and 448 (59%) hospitalizations, respectively, could have been prevented within the 2023-24 scenario. Increasing nirsevimab coverage to 90% by 1st October would avert up to 82% of hospitalizations in both subpopulations. In sensitivity analyses, if the catch-up group was immunized by 1st November, as per the Quebec 2024-25 roll-out, the NNI would increase from 53 to 69, consequently reducing averted hospitalizations from to 59% to 45% (Supplementary_Table 6).

Compared to hospitalization, about ten times as many nirsevimab immunizations are required to prevent one ICU admission at 556 and 537 for the at-birth and catch-up groups, respectively; however, based upon 2024-25 nirsevimab coverage, up to 60% of ICU admissions could be prevented and increasing to 90% coverage could prevent up to 76% (Table 2).

**Table 2:** Number needed to immunize to prevent an RSV hospitalization and ICU admission and potentially averted severe outcomes.

|  |  | Birth cohort |  |  |  |
| --- | --- | --- | --- | --- | --- |
|  |  | Born during 2023-24 season |  | Aged <6 months on October 1<sup>st</sup>, 2023 |  |
| Parameter | Data sources and calculations | Respiratory diagnosis | Bronchiolitis / bronchitis | Respiratory diagnosis | Bronchiolitis / bronchitis |
| **Number of respiratory hospitalizations** | Quebec hospitalization administrative database | 1566 | 863 | 1490 | 998 |
| **RSV positivity (%)** | Quebec hospital-based surveillance network | 65.4 | 81.3 | 51.0 | 67.3 |
| **Proportion of RSV hospitalizations with ICU admission (%)** | Quebec hospital-based surveillance network | 8.0 | 6.6 | 11.7 | 12.5 |
| **RSV hospitalizations** | Nb of hospitalizations * RSV positivity | 1024 | 702 | 760 | 672 |
| **RSV ICU admissions** | RSV hospitalizations * % of ICU admissions | 82 | 46 | 89 | 84 |
| **Births cohort (by eligible subpopulation)** | Institut de la statistique du Québec | 38250 | 38250 | 40050 | 40050 |
| **RSV hospitalization rate per 100000** | RSV hospitalizations / birth cohort | 2677.6 | 1834.3 | 1897.4 | 1677.0 |
| **RSV ICU admission rate per 100000** | RSV ICU admissions / birth cohort | 214.2 | 121.1 | 221.8 | 209.6 |
| **Effectiveness against hospitalization (95% CI)** | Quebec 2024-25 effectiveness | 91.5<br>(85.7–95.0) | 95.4<br>(89.4–98.0) | 91.5<br>(85.7–95.0) | 95.4<br>(89.4–98.0) |
| **Effectiveness against ICU admission (95% CI)** | Quebec 2024-25 effectiveness | 83.9<br>(39.3–95.7) | 83.9<br>(39.3–95.7) | 83.9<br>(39.3–95.7) | 83.9<br>(39.3–95.7) |
| **NNI to prevent one hospitalization** | 1 / (hospitalization rate * effectiveness and 95%CI) | 41<br>(39–44) | 57<br>(56–61) | 58<br>(51.3–56.8) | 63<br>(61–67) |
| **NNI to prevent one ICU admission** | 1 / (ICU admission rate * effectiveness and 95%CI) | 556<br>(488–1188) | 985<br>(863–2102) | 537<br>(471–1147) | 569<br>(498–1214) |
| **Nirsevimab coverage (%)** | 2024-25 immunization among test-neg controls | 79.2 | 79.2 | 64.4 | 64.4 |
| **(Potentially) averted hospitalizations** | (Coverage * Birth cohort) / NNI | 742 | 530 | 448 | 413 |
| **Proportion of hospitalizations averted (%)** | Averted hospitalizations / RSV hospitalizations | 72.5 | 75.6 | 58.9 | 61.4 |
| **(Potentially) averted ICU admissions** | (Coverage * Birth cohort) / NNI | 54 | 31 | 48 | 45 |
| **Proportion of ICU admissions averted (%)** | Averted ICU admissions / RSV ICU admissions | 66.5 | 66.5 | 54.0 | 54.0 |

Note: Scenario considering 2023-24 season between October 1<sup>st</sup>, 2023, and March 31<sup>st</sup>, 2024, and infants aged <6 months on October 1<sup>st</sup>, 2023, already immunized by the start of the season (October 1<sup>st</sup>)
Abbreviations: CI, confidence interval; ICU, intensive care unit; Nb, number; NNI, number needed to immunize; RSV, respiratory syncytial virus; test-neg, test-negative

## DISCUSSION

In this first population-based evaluation of universal nirsevimab immunization from Canada, we report substantial protection with effectiveness exceeding 85% against RSV-associated ER consultation, hospitalization and ICU admission. We estimate between 40 and 60 immunizations among newborns during the season and children <6 months at the start of the season, respectively, to prevent one hospitalization. Given such high effectiveness, ultimate program impact depends upon the level and timeliness of nirsevimab coverage achieved. We estimate two-thirds of RSV hospitalizations and 60% of ICU admissions could be prevented based upon 2024-25 coverage in Quebec but increasing the latter to 90% could prevent more than three-quarters of both outcomes. Conversely, delaying catch-up immunization by even a month could decrease averted hospitalizations by about 15%.

Our observed effectiveness of 89% (95%CI, 84%-92%) against RSV-associated hospitalization is consistent with observational data published to date and with efficacy results from randomized controlled trials (RCT). A recently published systematic review summarizing data from the USA, France, Italy, Luxembourg and Spain reported a pooled estimate of nirsevimab effectiveness of 83% against hospitalization [5], whereas pooled efficacy from RCTs is 81% [17]. Our estimated nirsevimab effectiveness of 88% (95%CI, 58%-97%) against RSV-associated ICU admissions and 86% (95%CI, 82%-90%) against RSV-associated ER consultations also aligns with published pooled effectiveness (81%) and efficacy (90%) data [5,17]. These results suggest potentially greater protection against hospitalization offered by nirsevimab versus maternal RSVpreF vaccination, with effectiveness of the latter estimated in Argentina, the first country to implement a maternal RSV vaccination program, to be 71% against infant RSV hospitalization during the first six months [18].

Although preterm infants were included in RCTs [19], nirsevimab effectiveness stratified for pre-term and other high-risk groups has not been estimated. Ours is the first study to estimate effectiveness against hospitalizations for preterm infants (93%) and children with high-risk chronic diseases (79%). While there are other considerations (e.g., economic), our findings of substantial benefit among children with or without risk factors for severe disease reinforce the impact of a universal program inclusive of healthy-term children, the latter estimated responsible for more than 80% of RSV-associated hospitalizations notwithstanding the highest individual risk of severe outcome among preterm infants and/or those with comorbidity [20].

Data on the duration of nirsevimab-induced protection are sparse. A few studies to date have suggested progressive decline over the first 5 months after administration [21,22]. A systematic review reported decrease in effectiveness with the study follow-up time, lower when the observation period was ≥150 days [23]. Our results suggest that nirsevimab effectiveness is maintained over the course of a season, or at least during the typical peak period in the northern hemisphere, but we also observed lower point estimates beyond four months post-administration. The low number of events during the last 10 weeks of the study period led to imprecision in effectiveness estimates among children immunized 12 to 20 weeks earlier. Bias due to the depletion of susceptibles could also explain an apparent pattern of waning: given high infant attack rates (exceeding 50%) [24], the proportion of non-immunized children being protected through infection-induced immunity will increase over the course of the season. If waning of protection at the tail end of the season is confirmed, this potential downside of early administration should be considered when establishing program launch dates.

RSV exerts important pressure on healthcare systems during the winter season, with the burden of hospitalizations and ICU admissions being especially high in infants aged <6 months [25]. In this first roll-out season in Quebec, nirsevimab potentially reduced 2024-2025 hospital admissions for RSV-associated respiratory diagnosis by 60% (72% in the at-birth and 45% in the catch-up groups). This resulted in a potential absolute reduction of 1075 hospitalizations with an acute respiratory infection among a birth cohort comprising just 86,000 infants (or 1,250 per 100,000 infants). The proportion of averted RSV-associated hospitalizations depends on timing of the immunization campaign and the start of the RSV season, with RSV seasonality suggesting that, optimally, catch-up immunization should be completed by October for maximal impact. Our estimated NNI was 40 for the at-birth group and 69 for the catch-up group if immunized by November, and 41 and 58, respectively, if immunized by October. Two Spanish studies reported NNI to prevent one hospitalization for at-birth immunization of 15 (from October to December) and 41 (from October to March) and at 71 and 90 for catch-up immunization [26,27]. Similarly, the NNI was estimated at 68 for infants aged <12 months at the start of the season using data from a multicenter RCT [17].

Our study has some limitations. Administrative databases for the 2024-25 season were incomplete at the time of our analysis with some hospitalizations still lacking a documented diagnosis and a proportion of recently born infants without a unique identifier as required to merge databases. Although this should not have compromised the validity of our effectiveness estimates, it reduced the available sample size contributing to imprecision. Also, some eligibility criteria for nirsevimab included a notion of clinical impact (i.e. cardiopathy with significant hemodynamic impact) which could not be adequately captured by ICD codes. Some included children may then have been not eligible for nirsevimab. Finally, estimations of NNI and averted hospitalizations depend on the RSV season and the cumulative hospitalization rates. We used the 2023-24 season, as data completeness allowed estimation of provincial rates and nirsevimab uptake was minimal. Although following similar seasonality, Quebec hospitalization surveillance showed that the 2024-25 RSV season was more intense than 2023-24 among age-groups not eligible for RSV immunization [28]. Our estimates of averted hospitalizations may therefore be conservative. RSV burden may be underestimated when based on administrative databases [29]. We overcame this limitation using RSV positivity from a surveillance network with systematic testing for respiratory admissions.

In conclusion, nirsevimab immunization provides substantial benefit against RSV-associated ER consultations, hospitalizations and ICU admissions to both healthy-term infants and high-risk groups. Maximizing coverage and impact through early-season program launch must take into account end-of-season durability of nirsevimab protection, the latter warranting further investigation.

## Acknowledgments

The authors thank Rémi Gagné, Pierre-Luc Trépanier, Marie-Claude Boisclair, Jonathan Phimmasone and Catherine Guimond (Institut national de santé publique du Québec) for their help with database preparedness, Quebec hospital-based surveillance data analysis and selection of ICD-10 codes for identification of chronic diseases.

## Author contributions

All the authors had full access to all the data in the study and take responsibility for the integrity of the data and the accuracy of the data analysis. Concept and design: S.C., R.G. and D.T. Acquisition, analysis, or interpretation of data: All authors. Drafting of the manuscript: S.C, J.P. and M.P. Critical revision of the manuscript for important intellectual content: All authors. Statistical analyses: M.O. and S.C. Supervision: J.P. and S.C.

## Data availability

Unpublished individual data are not available because the provincial registries used in this study are a property of the “Ministère de la Santé et des Services sociaux du Québec”. Data access to researchers who fulfill local requirements was authorized under the Health and Social Services Information Act. Aggregate data are available within the manuscript and the supplementary material.

## References

1. Suh M, Movva N, Jiang X, et al. Respiratory Syncytial Virus Is the Leading Cause of United States Infant Hospitalizations, 2009-2019: A Study of the National (Nationwide) Inpatient Sample. J Infect Dis 2022; 226:S154–S163.

2. Fitzpatrick T, Buchan SA, Mahant S, et al. Pediatric Respiratory Syncytial Virus Hospitalizations, 2017-2023. JAMA Netw Open 2024; 7:e2416077.

3. Drysdale SB, Cathie K, Flamein F, et al. Nirsevimab for Prevention of Hospitalizations Due to RSV in Infants. N Engl J Med 2023; 389:2425–2435.

4. Muller WJ, Madhi SA, Seoane Nuñez B, et al. Nirsevimab for Prevention of RSV in Term and Late-Preterm Infants. N Engl J Med 2023; 388:1533–1534.

5. Sumsuzzman DM, Wang Z, Langley JM, Moghadas SM. Real-world effectiveness of nirsevimab against respiratory syncytial virus disease in infants: a systematic review and meta-analysis. Lancet Child Adolesc Health 2025; 9:393–403.

6. Torres JP, Sauré D, Goic M, et al. Effectiveness and impact of nirsevimab in Chile during the first season of a national immunisation strategy against RSV (NIRSE-CL): a retrospective observational study. The Lancet Infectious Diseases 2025; :S1473309925002336.

7. Assad Z, Papenburg J, Ouldali N. Nirsevimab-a breakthrough in respiratory syncytial virus bronchiolitis. Lancet Child Adolesc Health 2024; :S2352-4642(24)00228–1.

8. Sanofi. Health Canada approves BEYFORTUS^TM^ (nirsevimab) for the prevention of RSV disease in infants. 2023. Available at: https://sanoficanada.mediaroom.com/2023-04-24-Health-Canada-approves-BEYFORTUS-TM-nirsevimab-for-the-prevention-of-RSV-disease-in-infants. Accessed 9 October 2024.

9. National Advisory Committee on Immunization (NACI), Killikelly A, Siu W, Brousseau N. Summary of the National Advisory Committee on Immunization (NACI) Statement on the Prevention of Respiratory Syncytial Virus (RSV) in Infants. CCDR 2025; 51:113–118.

10. Ministère de la Santé et des Services Sociaux du Québec. Ac-VRS : anticorps monoclonal contre le virus respiratoire syncytial (VRS). 2024. Available at: https://www.msss.gouv.qc.ca/professionnels/vaccination/piq-immunoglobulines/ac-vrs-anticorps-monoclonal-contre-virus-respiratoire-syncytial/. Accessed 15 March 2025.

11. Institut national d’excellence en santé et en services sociaux. Extract notice to the minister: Beyfortus (VRS). 2024. Available at: https://www.inesss.qc.ca/thematiques/medicaments/medicaments-evaluation-aux-fins-dinscription/extrait-davis-au-ministre/beyfortus-vrs-6995.html. Accessed 15 March 2025.

12. Paes B, Brown V, Courtney E, et al. Optimal implementation of an Ontario nirsevimab program for respiratory syncytial virus (RSV) prophylaxis: Recommendations from a provincial RSV expert panel. Human Vaccines & Immunotherapeutics 2024; 20. Available at: https://www.tandfonline.com/doi/full/10.1080/21645515.2024.2429236. Accessed 22 July 2025.

13. Government of Ontario. Respiratory Syncytial Virus (RSV) prevention programs. 2024. Available at: https://www.ontario.ca/page/respiratory-syncytial-virus-rsv-prevention-programs#:~:text=Ontario%20expanded%20the%20high%2Drisk,in%20infants%20and%20young%20children. Accessed 15 March 2025.

14. Ministère de la Santé et des Services sociaux’du Québec. VRS: vaccin contre le virus respiratoire syncitial. 2024. Available at: https://www.msss.gouv.qc.ca/professionnels/vaccination/piq-vaccins/vrs-vaccin-contre-virus-respiratoire-syncytial/. Accessed 15 March 2025.

15. Gilca R, Amini R, Carazo S, et al. The Changing Landscape of Respiratory Viruses Contributing to Hospitalizations in Quebec, Canada: Results From an Active Hospital-Based Surveillance Study. JMIR Public Health Surveill 2024; 10:e40792.

16. Institut de la statistique du Québec. Population et démographie. Population par année d’âge et par sexe, Québec. Available at: http://www.bdso.gouv.qc.ca/pls/ken/ken213_afich_tabl.page_tabl?p_iden_tran=REPERDUKTZY49-120149785127’zH2&p_lang=2&p_m_o=ISQ&p_id_ss_domn=986&p_id_raprt=697. Accessed 7 October 2019.

17. Jones JM, Fleming-Dutra KE, Prill MM, et al. Use of Nirsevimab for the Prevention of Respiratory Syncytial Virus Disease Among Infants and Young Children: Recommendations of the Advisory Committee on Immunization Practices — United States, 2023. MMWR Morb Mortal Wkly Rep 2023; 72:920–925.

18. Pérez Marc G, Vizzotti C, Fell DB, et al. Real-world effectiveness of RSVpreF vaccination during pregnancy against RSV-associated lower respiratory tract disease leading to hospitalisation in infants during the 2024 RSV season in Argentina (BERNI study): a multicentre, retrospective, test-negative, case–control study. Lancet Infect Dis 2025; :S1473309925001562.

19. Simões EAF, Madhi SA, Muller WJ, et al. Efficacy of nirsevimab against respiratory syncytial virus lower respiratory tract infections in preterm and term infants, and pharmacokinetic extrapolation to infants with congenital heart disease and chronic lung disease: a pooled analysis of randomised controlled trials. Lancet Child Adolesc Health 2023; 7:180–189.

20. Pisesky A, Benchimol EI, Wong CA, et al. Incidence of Hospitalization for Respiratory Syncytial Virus Infection amongst Children in Ontario, Canada: A Population-Based Study Using Validated Health Administrative Data. PLoS ONE 2016; 11:e0150416.

21. Barbas Del Buey JF, Íñigo Martínez J, Gutiérrez Rodríguez MÁ, et al. The effectiveness of nirsevimab in reducing the burden of disease due to respiratory syncytial virus (RSV) infection over time in the Madrid region (Spain): a prospective population-based cohort study. Front Public Health 2024; 12:1441786.

22. Xu H, Aparicio C, Wats A, et al. Estimated Effectiveness of Nirsevimab Against Respiratory Syncytial Virus. JAMA Netw Open 2025; 8:e250380.

23. Riccò M, Cascio A, Corrado S, et al. Impact of Nirsevimab Immunization on Pediatric Hospitalization Rates: A Systematic Review and Meta-Analysis (2024). Vaccines 2024; 12:640.

24. Rosas-Salazar C, Chirkova T, Gebretsadik T, et al. Respiratory syncytial virus infection during infancy and asthma during childhood in the USA (INSPIRE): a population-based, prospective birth cohort study. The Lancet 2023; 401:1669–1680.

25. Bourdeau M, Vadlamudi NK, Bastien N, et al. Pediatric RSV-Associated Hospitalizations Before and During the COVID-19 Pandemic. JAMA Netw Open 2023; 6:e2336863.

26. Ezpeleta G, Navascués A, Viguria N, et al. Effectiveness of Nirsevimab Immunoprophylaxis Administered at Birth to Prevent Infant Hospitalisation for Respiratory Syncytial Virus Infection: A Population-Based Cohort Study. Vaccines 2024; 12:383.

27. Pastor-Barriuso R, Núñez O, Monge S, the Nirsevimab Effectiveness Study Collaborators. Infants needed to immunise with nirsevimab to prevent one RSV hospitalisation, Spain, 2023/24 season. Euro Surveill 2025; 30. Available at: https://www.eurosurveillance.org/content/10.2807/1560-7917.ES.2025.30.6.2500040. Accessed 24 June 2025.

28. Ministère de la Santé et des Services sociaux du Québec. Infections par le virus respiratoire syncitial (VRS). 2025. Available at: https://www.msss.gouv.qc.ca/professionnels/maladies-infectieuses/infections-virus-respiratoire-syncytial-vrs/suivi-activite-vrs/. Accessed 20 May 2025.

29. Jollivet O, Urchueguía-Fornes A, Chung-Delgado K, et al. Respiratory syncytial virus hospitalisation burden in children below 18 years in six European countries (2016-2023) pre- and post-COVID-19 pandemic. Int J Infect Dis 2025; 155:107903.

