## Supplemental material for "Nirsevimab effectiveness, number needed to immunize and impact on severe RSV outcomes in preterm, high-risk and healthy-term infants, Quebec, Canada"

### Supplementary Appendix

Supplementary Table 1. ICD-10 codes for high-risk chronic conditions………….....…….2

Supplementary Table 2. ICD-10 codes for respiratory and bronchiolitis / bronchitis diagnosis ………………………………………..……………………….…………………..3

Supplementary Table 3. Odds ratios of immunization and nirsevimab effectiveness against ER consultation by outcome definition, eligible subpopulation and age group.....4

Supplementary Table 4. Odds ratios of immunization and nirsevimab effectiveness against hospitalization by outcome definition, eligible subpopulation and age group...….5

Supplementary Table 5. Odds ratios of immunization and nirsevimab effectiveness against ICU admission by outcome definition, eligible subpopulation and age group.......6

Supplementary Table 6: Number needed to immunize to prevent an RSV hospitalization and potentially averted hospitalizations during the 2023-24 season, sensitivity analyses for immunization delayed by one month (by 1st November)……………………….…….7

### Supplementary Table 1. ICD-10 codes for high-risk chronic conditions

| Chronic condition | ICD-10 codes |
|---|---|
| Bronchopulmonary dysplasia | P23-P2699; Q32-Q3499 |
| Chronic pulmonary disease | J40-J4799; J84-J8499; J98-J9999; P27-P2899 |
| Congenital heart disease or cardiomyopathy | P29-P2999; Q20-Q2899; I31-I3199; I34-I3799; I42-I4299 |
| Moderate or severe pulmonary arterial hypertension | I27-I2899 |
| Down syndrome | Q90-Q9099 |
| Cystic fibrosis | E84-E8499 |
| Neuromuscular disorder | D43-D4399; G11-G1399; G24-G2499; G31-G3299; G37-G3799; G40-G4099; G70-G7199; K449; P91-P9199; Q00-Q0799; Q31-Q3299; Q34-Q3999; Q79-Q7999; Q87-Q8799; Q89-Q899; R29-R2999 |
| Bone marrow, stem cell or solid organ transplant | Z94-Z949 |
| Asthma, emphysema | J4590; J439 |
| Prematurity | P07-P079 |

**Supplementary Table 2. ICD-10 codes for respiratory and bronchiolitis / bronchitis diagnosis**

| Diagnosis | ICD-10 codes |
|---|---|
| Bronchiolitis or Bronchitis | J20.0-J20.9; J21*; J21.0-J21.9; J22-J22.9; J40; J980 |
| Pneumonia | J10.0; J12.0-J12.9; J13; J14; J15.0-J15.9; J16.0-J16.8; J17; J18.0-J18.0 |
| Other diagnosis for acute respiratory infection | A73.0-A73.9; A48.1; B25.0; B34.2-B34.9; B44.0; B77.81; B97.0-B97.88; J00-J069; J09X1-J09X9; J10.1-J10.89; J80-J809; J96-J9692; R042; R05-R07.1; R09-R09.2; R50.9; U04-U04.99; U07.1-U07.2; B05.0-B06.9; B08.2-B08.5; B33.8; B34.0-B34.9; B37.0-B37.1; H65.0-H67.8; R50.8-R50.9; R56.01-R56.88; J45.00-J45.91; J38.4-J39.9; J47; J69.0; J96.00-J96.99; J98.0-J98.9 |

### Supplementary Table 3: Odds ratios of immunization and nirsevimab effectiveness against ER consultation by outcome definition, eligible subpopulation and age group

| | Cases | Controls | Unadjusted OR (95% CI) | Adjusted OR[a] (95% CI) | Effectiveness[b] (95% CI) |
|---|---|---|---|---|---|
| **By outcome definition, with positive RSV test** | | | | | |
| ER consultation (main analysis) | | | | | |
| Non-immunized | 599 | 1239 | Reference | Reference | Reference |
| Nirsevimab | 69 | 1265 | 0.11 (0.09–0.15) | 0.14 (0.10–0.19) | 86% (82–90) |
| Respiratory ER consultation | | | | | |
| Non-immunized | 587 | 1148 | Reference | Reference | Reference |
| Nirsevimab | 68 | 1188 | 0.11 (0.09–0.15) | 0.14 (0.10–0.18) | 87% (82–90) |
| ER consultation for bronchiolitis / bronchitis | | | | | |
| Non-immunized | 176 | 97 | Reference | Reference | Reference |
| Nirsevimab | 20 | 72 | 0.15 (0.09–0.27) | 0.11 (0.05–0.24) | 89% (76–95) |
| **By eligible subpopulation** [c] | | | | | |
| Healthy term children | | | | | |
| Non-immunized | 424 | 885 | Reference | Reference | Reference |
| Nirsevimab | 52 | 978 | 0.11 (0.08–0.15) | 0.14 (0.10–0.20) | 86% (80–90) |
| Children with chronic diseases | | | | | |
| Non-immunized | 123 | 278 | Reference | Reference | Reference |
| Nirsevimab | 12 | 172 | 0.16 (0.09–0.29) | 0.14 (0.07–0.30) | 86% (70–93) |
| Preterm children | | | | | |
| Non-immunized | 59 | 93 | Reference | Reference | Reference |
| Nirsevimab | 6 | 155 | 0.06 (0.03–0.15) | 0.03 (0.01–0.09) | 97% (91–99) |
| **By age group** [c,d] | | | | | |
| Newborn or <6 months | | | | | |
| Non-immunized | 501 | 1015 | Reference | Reference | Reference |
| Nirsevimab | 64 | 1186 | 0.11 (0.08–0.14) | 0.13 (0.09–0.17) | 87% (83–91) |
| Newborn during season | | | | | |
| Non-immunized | 30 | 132 | Reference | Reference | Reference |
| Nirsevimab | 21 | 492 | 0.19 (0.10–0.34) | 0.17 (0.08–0.35) | 83% (65–92) |
| <6 months | | | | | |
| Non-immunized | 471 | 883 | Reference | Reference | Reference |
| Nirsevimab | 43 | 694 | 0.12 (0.08–0.16) | 0.11 (0.08–0.16) | 89% (84–92) |
| 6-18 months | | | | | |
| Non-immunized | 98 | 224 | Reference | Reference | Reference |
| Nirsevimab | 5 | 79 | 0.15 (0.06–0.37) | 0.09 (0.03–0.27) | 91% (74–97) |

[a] Adjusted for age group, sex, region of residence, prematurity, presence of comorbidities, two-week period of testing

[b] Nirsevimab effectiveness calculated as 1 – adjusted OR

[c] Outcome defined as hospitalization with positive RSV test

[d] Newborns during the season or age on October 1st, 2024

Abbreviations: CI, confidence interval; ER, emergency room; OR, odds ratio; RSV, respiratory syncytial virus

**Supplementary Table 4: Odds ratios of immunization and nirsevimab effectiveness against hospitalization by outcome definition, eligible subpopulation and age group**

| | Cases | Controls | Unadjusted OR (95% CI) | Adjusted OR[a] (95% CI) | Effectiveness[b] (95% CI) |
|---|---|---|---|---|---|
| **By outcome definition, with positive RSV test** | | | | | |
| Hospitalization (main analysis) | | | | | |
| Non-immunized | 494 | 582 | Reference | Reference | Reference |
| Nirsevimab | 55 | 627 | 0.10 (0.08–0.14) | 0.11 (0.08–0.16) | 89% (84–92) |
| Respiratory hospitalization | | | | | |
| Non-immunized | 417 | 271 | Reference | Reference | Reference |
| Nirsevimab | 34 | 220 | 0.10 (0.07–0.15) | 0.08 (0.05–0.12) | 93% (88–95) |
| Hospitalization for bronchiolitis / bronchitis | | | | | |
| Non-immunized | 357 | 92 | Reference | Reference | Reference |
| Nirsevimab | 26 | 74 | 0.09 (0.06–0.15) | 0.04 (0.02–0.09) | 96% (91–98) |
| **By eligible subpopulation** [c] | | | | | |
| Healthy term children | | | | | |
| Non-immunized | 322 | 322 | Reference | Reference | Reference |
| Nirsevimab | 29 | 379 | 0.08 (0.05–0.12) | 0.10 (0.06–0.16) | 90% (84–94) |
| Children with chronic diseases | | | | | |
| Non-immunized | 107 | 218 | Reference | Reference | Reference |
| Nirsevimab | 18 | 184 | 0.20 (0.12–0.34) | 0.21 (0.11–0.40) | 79% (60–89) |
| Preterm children | | | | | |
| Non-immunized | 76 | 59 | Reference | Reference | Reference |
| Nirsevimab | 14 | 125 | 0.09 (0.05–0.17) | 0.07 (0.03–0.17) | 93% (83–97) |
| **By age group** [c,d] | | | | | |
| Newborn or <6 months | | | | | |
| Non-immunized | 405 | 416 | Reference | Reference | Reference |
| Nirsevimab | 49 | 540 | 0.09 (0.07–0.13) | 0.11 (0.08–0.17) | 89% (83–92) |
| Newborn during season | | | | | |
| Non-immunized | 32 | 51 | Reference | Reference | Reference |
| Nirsevimab | 16 | 199 | 0.13 (0.07–0.25) | 0.16 (0.06–0.42) | 84% (58–94) |
| <6 months | | | | | |
| Non-immunized | 373 | 365 | Reference | Reference | Reference |
| Nirsevimab | 33 | 341 | 0.10 (0.06–0.14) | 0.09 (0.06–0.15) | 91% (85–94) |
| 6-18 months | | | | | |
| Non-immunized | 89 | 146 | Reference | Reference | Reference |
| Nirsevimab | 6 | 69 | 0.14 (0.06–0.34) | 0.10 (0.04–0.25) | 91% (75–96) |

[a] Adjusted for age group, sex, region of residence, prematurity, presence of comorbidities, two-week period of testing

[b] Nirsevimab effectiveness calculated as 1 – adjusted OR

[c] Outcome defined as hospitalization with positive RSV test

[d] Newborns during the season or age on October 1st, 2024

Abbreviations: CI, confidence interval; OR, odds ratio; RSV, respiratory syncytial virus

### Supplementary Table 5: Odds ratios of immunization and nirsevimab effectiveness against ICU admission by outcome definition, eligible subpopulation and age group

| | Cases | Controls | Unadjusted OR (95% CI) | Adjusted OR[a] (95% CI) | Effectiveness[b] (95% CI) |
|---|---|---|---|---|---|
| **By outcome definition, with positive RSV test** | | | | | |
| ICU admission (main analysis) | | | | | |
| Non-immunized | 31 | 582 | Reference | Reference | Reference |
| Nirsevimab | 3 | 627 | 0.09 (0.03–0.30) | 0.12 (0.03–0.42) | 88% (58–97) |
| **By eligible subpopulation** [c] | | | | | |
| Healthy term children | | | | | |
| Non-immunized | 21 | 322 | Reference | Reference | Reference |
| Nirsevimab | 2 | 379 | 0.08 (0.02–0.35) | 0.12 (0.02–0.59) | 88% (42–98) |
| Children with chronic diseases | | | | | |
| Non-immunized | 8 | 218 | Reference | | |
| Nirsevimab | 1 | 184 | 0.38 (0.04–3.31) | NE | NE |
| Preterm children | | | | | |
| Non-immunized | 4 | 59 | | | |
| Nirsevimab | 0 | 125 | NE | NE | NE |
| **By age group** [c,d] | | | | | |
| Newborn or <6 months | | | | | |
| Non-immunized | 25 | 416 | Reference | Reference | Reference |
| Nirsevimab | 3 | 540 | 0.09 (0.03–0.31) | 0.16 (0.04–0.61) | 84% (39–96) |
| Newborn during season | | | | | |
| Non-immunized | 0 | 51 | | | |
| Nirsevimab | 1 | 199 | NE | NE | NE |
| <6 months | | | | | |
| Non-immunized | 25 | 365 | Reference | | |
| Nirsevimab | 2 | 341 | 0.09 (0.02–0.36) | NE | NE |
| 6-18 months | | | | | |
| Non-immunized | 6 | 166 | | | |
| Nirsevimab | 0 | 87 | NE | NE | NE |

[a] Adjusted for age group, sex, region of residence, prematurity, presence of comorbidities, two-week period of testing

[b] Nirsevimab effectiveness calculated as 1 – adjusted OR

[c] Outcome defined as hospitalization with positive RSV test

[d] Newborns during the season or age on October 1st, 2024

Abbreviations: CI, confidence interval; NE, not estimable; ICU, intensive care unit; OR, odds ratio; RSV, respiratory syncytial virus

### Supplementary Table 6: Number needed to immunize to prevent an RSV hospitalization and ICU admission and potentially averted severe outcomes for immunization delayed by one month (by 1st November)

| | | Birth cohort | | | |
|---|---|---|---|---|---|
| | | Born during 2023-24 season | | Aged <6 months on October 1st, 2023 | |
| **Parameter** | **Data sources and calculations** | Respiratory diagnosis | Bronchiolitis / bronchitis | Respiratory diagnosis | Bronchiolitis / bronchitis |
| **Number of respiratory hospitalizations** | Quebec hospitalization administrative database | 1548 | 851 | 1150 | 803 |
| **RSV positivity** (%) | Quebec hospital-based surveillance network | 65.4 | 81.3 | 50.4 | 66.5 |
| **Proportion of RSV hospitalizations with ICU admission** (%) | Quebec hospital-based surveillance network | 8.0 | 6.6 | 10.8 | 11.3 |
| **RSV hospitalizations** | Nb of hospitalizations * RSV positivity | 1012 | 692 | 580 | 534 |
| **RSV ICU admissions** | RSV hospitalizations * % of ICU admissions | 81 | 46 | 62 | 60 |
| **Births cohort (by eligible subpopulation)** | Institut de la statistique du Québec | 38250 | 38250 | 40050 | 40050 |
| **RSV hospitalization rate per 100000** | RSV hospitalizations / birth cohort | 2646.8 | 1808.8 | 1447.2 | 1333.3 |
| **RSV ICU admission rate per 100000** | RSV ICU admissions / birth cohort | 211.7 | 119.4 | 155.9 | 150.7 |
| **Effectiveness against hospitalization** (95% CI) | Quebec 2024-25 effectiveness | 91.5 (85.7–95.0) | 95.4 (89.4–98.0) | 91.5 (85.7–95.0) | 95.4 (89.4–98.0) |
| **Effectiveness against ICU admission** (95% CI) | Quebec 2024-25 effectiveness | 83.9 (39.3–95.7) | 83.9 (39.3–95.7) | 83.9 (39.3–95.7) | 83.9 (39.3–95.7) |
| **NNI to prevent one hospitalization** | 1 / (hospitalization rate * effectiveness and 95%CI) | 41 (40–44) | 59 (56–62) | 76 (73–81) | 79 (77–84) |
| **NNI to prevent one ICU admission** | 1 / (ICU admission rate * effectiveness and 95%CI) | 563 (493–1202) | 998 (875–2131) | 765 (670–1633) | 791 (694–1689) |
| **Nirsevimab coverage** (%) | 2024-25 immunization among test-neg controls | 79.2 | 79.2 | 64.4 | 64.4 |
| **(Potentially) averted hospitalizations** | (Coverage * Birth cohort) / NNI | 734 | 518 | 342 | 325 |
| **Proportion of hospitalizations averted** (%) | Averted hospitalizations / RSV hospitalizations Oct-March | 71.7 | 73.8 | 58.9 | 61.4 |
| **(Potentially) averted ICU admissions** | (Coverage * Birth cohort) / NNI | 54 | 30 | 34 | 33 |
| **Proportion of ICU admissions averted** (%) | Averted ICU admissions / RSV ICU admissions Oct-March | 65.7 | 65.5 | 44.9 | 48.4 |

Note: Scenario considering 2023-24 season between October 1st, 2023, and March 31st, 2024, and infants aged <6 months on October 1st, 2023, immunized only by November 1st, 2023

Abbreviations: CI, confidence interval; ICU, intensive care unit; Nb, number; NNI, number needed to immunize; RSV, respiratory syncytial virus; test-neg, test-negative
